# Comparison of Flow Ratio Derived From Intravascular Ultrasound with Coronary Angiography in Complex Coronary Artery Lesions: Correlation with Fractional Flow Reserve

**DOI:** 10.1101/2025.04.16.25325975

**Authors:** Rende Xu, Zhe Wang, Congcong Pan, Wei Gao, Leilei Ma, Yizhe Wu, Zhangwei Chen, Yang Lu, Yong Dong, Wei Yu, Shengxian Tu, Chenguang Li, Junbo Ge

**Author notes:** Rende Xu and Zhe Wang contributed equally to this work. **Conflict of interest statement:** All the authors declare that there are no conflicts of interest. **Corresponding author**: Dr. Chenguang Li, Department of Cardiology, Zhongshan Hospital, Fudan University, 180 Fenglin Road, Shanghai 200032, China.; Dr. Wei Yu, Room 123, Med-X Research Institute, Shanghai Jiao Tong University, No. 1954, Hua Shan Rd, Shanghai 200030, China.

## Abstract

**Background:** Accurate assessment of coronary artery disease is essential for guiding clinical decision-making, particularly in cases involving complex coronary lesions. Fractional Flow Reserve (FFR) remains the reference standard for lesion-specific ischemia evaluation; however, it is invasive and requires pharmacologically induced hyperemia. Emerging non-invasive alternatives, including Quantitative Flow Ratio (QFR) derived from coronary angiography (CAG) and Ultrasonic Flow Ratio (UFR) derived from intravascular ultrasound (IVUS), offer promising diagnostic value. This study aimed to compare the diagnostic performance of UFR and QFR against FFR in the assessment of complex coronary lesions.

**Methods:** In this retrospective, multicenter study, 217 patients (220 vessels) with coronary artery lesions who underwent both intravascular ultrasound (IVUS) and FFR measurement were included. UFR was derived from IVUS imaging, while QFR was computed using coronary angiography (CAG) data. Correlation coefficients, agreement analyses, and diagnostic metrics including sensitivity and specificity were employed to evaluate the performance of UFR and QFR against FFR, with receiver operating characteristic (ROC) curve analysis used to assess diagnostic accuracy.

**Results:** UFR demonstrated a stronger correlation with FFR (r = 0.79, *p* < 0.001) compared to QFR (r = 0.68, *p* < 0.001). Moreover, UFR exhibited superior diagnostic performance, with an area under the ROC curve (AUC) of 0.91, exceeding that of QFR (AUC = 0.86). In subsets of complex lesions—specifically diffuse, bifurcation, and heavily calcified lesions— UFR consistently outperformed QFR, with the most pronounced difference observed in bifurcation and calcified lesions where QFR accuracy was significantly reduced (72.5% vs. 86.8%, *p* = 0.001).

**Conclusion:** UFR provides enhanced diagnostic accuracy compared to QFR for complex coronary lesions and represents a reliable, non-invasive alternative to FFR, particularly in challenging anatomical scenarios such as bifurcation and heavily calcified lesions. The clinical integration of UFR may reduce the reliance on invasive FFR measurements while preserving diagnostic precision.

## INTRODUCTION

Coronary artery disease (CAD) remains a significant global health concern, necessitating precise diagnostic strategies for optimal patient management ^1^. Fractional flow reserve (FFR), defined as the distal-to-proximal pressure ratio across a coronary stenosis during maximal hyperemia, is the gold standard for detecting ischemia-causing stenoses and guiding revascularization decisions ^2,3^. However, the widespread use of FFR is limited by its invasive nature and the requirement for pharmacological hyperemia, which can be cumbersome and uncomfortable for patients. Consequently, there is growing interest in developing and validating non-invasive or minimally invasive alternatives derived from anatomical imaging data ^4,5^.

Quantitative flow ratio (QFR) is a promising non-invasive method derived from coronary angiography (CAG). It has been extensively investigated, showing high diagnostic performance in identifying hemodynamically significant lesions. However, CAG-based assessments have inherent limitations in terms of capturing detailed vessel and plaque morphology ^6,7^. Intravascular ultrasound (IVUS) offers superior resolution for evaluating vessel lumen size and characterizing plaque, which has led to the development of Ultrasonic flow ratio (UFR), an IVUS-based approach that integrates both anatomical and physiological evaluations into a single assessment ^8,9^.

While both QFR and UFR have demonstrated effectiveness, their relative performances in complex coronary lesions, such as diffuse, bifurcation, and heavily calcified lesions, are not yet fully understood. Complex lesions are associated with higher risks of visual-functional mismatches, complicating the assessment of lesion severity ^10,11^. In this real-world, multicenter study, we compare the diagnostic performance of QFR and UFR against the gold-standard FFR, with a particular focus on complex coronary lesions. This evaluation aims to determine the advantages and limitations of each modality and explore their utility in the clinical decision-making process for complex CAD cases.

## METHODS

### Patient population

This study represents a multicenter, retrospective observational analysis conducted at Zhongshan Hospital in Shanghai, Shanghai 7th People’s Hospital, and Zhengzhou 7th People’s Hospital. It included a cohort of consecutive patients who underwent both IVUS imaging and FFR measurement on the same artery, spanning from July 2018 to April 2023. Participants had at least one target lesion with 30% to 90% diameter stenosis, as visually estimated via CAG. Exclusion criteria comprised: (1) insufficient quality of CAG or IVUS images for QFR or UFR calculations, (2) prior balloon predilatation or stent placement before FFR measurement or IVUS imaging, (3) incomplete IVUS pullback across the entire lesion segment, (4) presence of a severe myocardial bridge (≥30% systolic diameter stenosis) in the examined vessel, (5) history of previous coronary artery bypass grafting (CABG), and (6) left ventricular ejection fraction (LVEF) <35%. Written informed consent was obtained from all participants for the procedure and for the collection and analysis of data for research purposes. The study received approval from the institutional review boards at all three participating sites and was registered with ClinicalTrials.gov (NCT06322355).

### Invasive CAG, FFR and IVUS

Invasive CAG was conducted following standard clinical procedures, encompassing multiple projections of both the left and right coronary arteries. Prior to the FFR measurement and IVUS imaging, each patient was administered 5,000 IU of intravenous heparin and 0.2 mg of intracoronary nitroglycerin. The FFR was precisely measured using either an intracoronary pressure wire (Pressure Wire X; Abbott Vascular, Santa Clara, USA) or a rapid-exchange pressure microcatheter (Insight Lifetech, Shanghai, China) during peak hyperemia. This state of maximal hyperemia was achieved through the intravenous infusion of adenosine triphosphate (ATP) at a concentration of 150 μg/kg/min, administered via the forearm vein. Upon completion of the FFR assessment, the pressure wire or microcatheter was carefully retracted to the tip of the guiding catheter for a routine drift check. IVUS imaging was meticulously conducted using the iLab™ IVUS system, coupled with a 40-MHz OptiCross IVUS catheter (Boston Scientific, Fremont, USA), at a steady pullback speed of 0.5 mm/s.

### QFR and UFR measurements

QFR and UFR measurements were performed at an image core laboratory who was blinded from clinical data and wire-based FFR. QFR analysis was conducted offline using the AngioPlus software (Pulse Medical Imaging Technology, Shanghai, China), following protocols comparable to those in previous studies. In summary, the computation of QFR requires an optimal angiographic image. For this purpose, two angiographic images with at least a 25° difference in projection angles were uploaded into the AngioPlus system. This facilitated the three-dimensional reconstruction of the targeted vessel, excluding its side branches. The lumen contour was then automatically delineated using highly validated algorithms. The QFR computation relied on the contrast flow model, which integrates contrast flow velocity, adapted from modified TIMI frame counts. It’s important to note that these QFR computations were conducted by researchers who were not privy to the FFR results. A hemodynamically significant coronary stenosis is determined when the QFR is ≤0.80.

UFR computation was performed by an experienced analyst who was blinded to the FFR and QFR results using a prototype software package (IvusPlus prototype, Pulse Medical Imaging Technology, Shanghai, China). The detailed method for this calculation has been described before ^8^. A deep learning model, RefineNet10, was used to automatically identify and reconstruct artery images in 3D. Experienced analysts first outlined the IVUS images, then the model learned to identify different parts of the artery. Consecutive IVUS cross-sectional images were also used to understand blood flow patterns. If the model’s outlining was inaccurate, manual corrections were made. They also reconstructed and measured the areas of side branches of arteries. They then estimated the healthy size of the artery and calculated the blood flow based on a standard flow velocity. Finally, they calculated pressure drops at different points in the artery using a computational method that considers various factors like friction and sudden changes in flow.

A diffuse lesion was defined as a single stenotic segment with a length of ≥20 mm. A bifurcation lesion was characterized by coronary artery stenosis located near or involving the origin of a significant side branch with high functional importance. A heavily calcified lesion was identified when the length of calcification on both sides of the target lesion exceeded 50 mm, or if IVUS showed a calcium arc spanning more than 270°.

### Statistical analysis

Statistical analysis was performed using MedCalc software (version 20.217). Categorical variables were described as numbers and percentages. Continuous variables are reported as the mean ± standard deviation (SD). Correlation was evaluated using Pearson or Spearman correlation as appropriate. Bland-Altman plots and intraclass correlation coefficients (ICC) for the absolute values were used for assessing agreement between different continuous parameters. Comparison of the limits of agreement between QFR and UFR was performed by the F-test. A Wilcoxon signed-rank test or paired t-test was used for pairwise comparison as appropriate. Diagnostic performance was assessed using the area under the curve (AUC) by receiver operating characteristic (ROC) analysis. An area of 1.0 indicated perfect performance, while 0.5 indicated a performance that could not be differentiated from chance. All p-values reported are two-tailed. Statistical significance was set at 0.05.

## RESULTS

### Patient and lesion characteristics

In total, data from 248 vessels belonging to 242 patients were initially screened for inclusion. Of these, 28 vessels were excluded based on predefined exclusion criteria: 9 due to inadequate angiographic or IVUS image quality, 1 due to the presence of severely tortuous vessels, 10 due to incomplete IVUS pullback, 1 due to a significant myocardial bridge, 6 due to prior CABG, and 1 due to LVEF <35%. As a result, the final study cohort consisted of 217 patients and 220 coronary vessels (Figure 1). Among these, only 82 vessels (37.3%) met the criteria for optimal QFR image quality. The mean age of the patients was 63.4 ± 9.9 years, with 162 (74.7%) being male. Pertinent demographics and clinical characteristics are detailed in Table 1.

**Figure 1.**
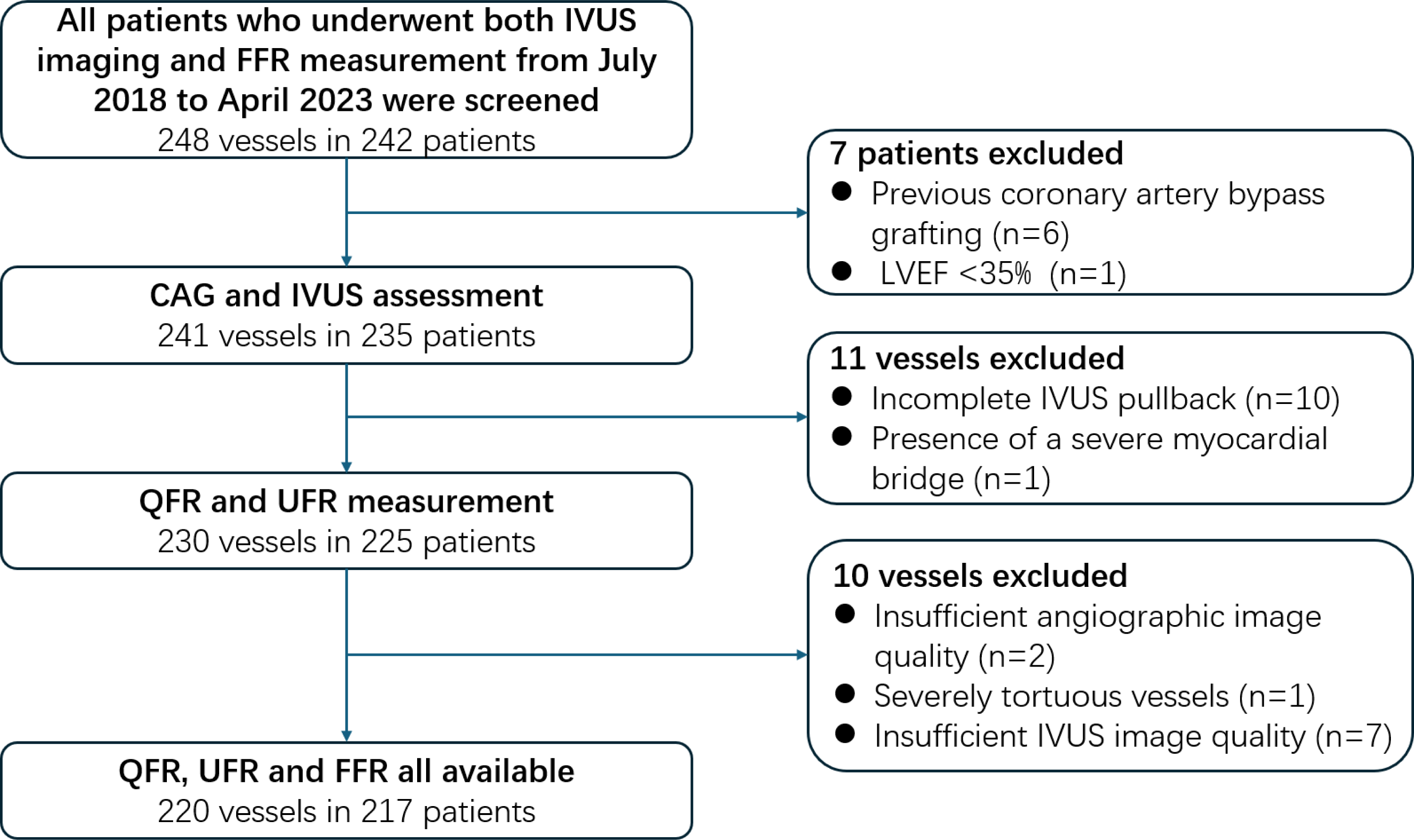
Study flowchart. IVUS, intravascular ultrasound; FFR, fractional flow reserve; CAG, coronary angiography; QFR, quantitative flow reserve; UFR, ultrasonic flow ratio; LVEF, left ventricular ejection fraction.

**Table 1.** Baseline Characteristics of Patients.

| Characteristics | Value |
| --- | --- |
| Demographic data |  |
| Age, y | 63.4 ± 9.9 |
| Male, n (%) | 162 (74.7) |
| Body mass index, kg/m<sup>2</sup> | 25.2 ± 3.0 |
| Cardiovascular risk factors |  |
| Hypertension, n (%) | 140 (64.5) |
| Dyslipidemia, n (%) | 50 (23.0) |
| Diabetes mellitus, n (%) | 63 (29.0) |
| Smoking, n (%) | 81 (37.3) |
| Familial history of premature CAD, n (%) | 13 (6.0) |
| History |  |
| Previous myocardial infarction, n (%) | 14 (6.5) |
| Previous PCI, n (%) | 56 (25.8) |
| Peripheral artery disease, n (%) | 5 (2.3) |
| Atrial fibrillation, n (%) | 6 (2.8) |
| Chronic kidney disease | 4 (1.8) |
| Medical therapy on admission |  |
| Aspirin, n (%) | 181 (83.4) |
| Clopidogrel/Ticagrelor, n (%) | 157 (72.4) |
| Statins, n (%) | 210 (96.8) |
| β-blockers, n (%) | 132 (60.8) |
| Calcium channel blockers, n (%) | 181 (83.4) |
| ACE inhibitors or AT-II receptor antagonist, n (%) | 111 (51.2) |
| Nitrates, n (%) | 74 (34.1) |
| UCG parameters |  |
| LVEF, % | 65.0 ± 4.3 |
| LVESd, mm | 29.6 ± 3.1 |
| LVEDd, mm | 46.5 ± 4.1 |

CAD indicates coronary artery disease; PCI, percutaneous coronary intervention; ACE, angiotensin converting enzyme; AT, angiotensin; UCG, ultrasoundcardiography; LVEF, left ventricular ejection fraction; LVESd, left ventricular end-systolic diameter; LVEDd, left ventricular end-diastolic diameter.

The left anterior descending artery was the most frequently examined vessel (84.6%), followed by the right coronary artery (10.9%) and the left circumflex artery (4.5%). Among the vessels included, 120 vessels had diffuse lesions, 75 had bifurcation lesions, and 91 had heavily calcified lesions. Quantitative coronary angiography (QCA) revealed an average percent diameter stenosis (%DS) of 42.7 ± 8.7%, with an average QFR of 0.81 ± 0.11. The average minimum lumen area (MLA) measured by IVUS was 3.2 ± 1.2 mm^2^, and the average UFR was 0.79 ± 0.10. Across all lesions, the mean FFR value was 0.79 ± 0.10, with a range of 0.55 to 0.99. Notably, 92 lesions (41.8%) exhibited FFR values falling within the range of 0.75 to 0.85. Detailed lesion characteristics are summarized in Table 2.

**Table 2.** Lesion Characteristic.

| Characteristics | Value |
| --- | --- |
| Native vessel disease, n (%) | 210 (95.5) |
| In-Stent restenosis, n (%) | 10 (4.5) |
| Vessel |  |
| LAD, n (%) | 186 (84.6) |
| LCx, n (%) | 10 (4.5) |
| RCA, n (%) | 24 (10.9) |
| Morphological characteristics |  |
| Diffuse lesion, n (%) | 120 (54.5) |
| Bifurcation lesion, n (%) | 75 (34.1) |
| Heavily calcified lesion, n (%) | 91 (41.4) |
| FFR based characteristics |  |
| FFR value | 0.79 ± 0.10 |
| 0.75 ≤ FFR ≤ 0.85, n (%) | 92 (41.8) |
| QCA/QFR based characteristics |  |
| Diameter stenosis, % | 42.7 ± 8.7 |
| Lesion length, mm | 31.1 ± 15.2 |
| Minimal lumen diameter, mm | 1.7 ± 0.5 |
| Minimal lumen area, mm<sup>2</sup> | 2.5 ± 1.3 |
| Reference vessel diameter, mm | 3.2 ± 2.5 |
| QFR value | 0.81 ± 0.11 |
| IVUS/UFR based characteristics |  |
| Lesion length, mm | 41.1 ± 19.2 |
| Minimal lumen area, mm<sup>2</sup> | 3.2 ± 1.2 |
| Maximal plaque burden, % | 73.2 ± 9.0 |
| Proximal reference area, mm<sup>2</sup> | 4.8 ± 1.3 |
| Distal reference area, mm<sup>2</sup> | 3.1 ± 0.8 |
| Plaque volume, mm<sup>3</sup> | 305.9 ± 173.6 |
| Lumen volume, mm<sup>3</sup> | 296.3 ± 165.5 |
| UFR value | 0.79 ± 0.10 |

LAD, left anterior descending artery; LCx, left circumflex artery; RCA, right coronary artery; FFR, fractional flow reserve; QCA, quantitative coronary angiography; QFR, quantitative flow reserve; IVUS, intravascular ultrasound; UFR, ultrasonic flow ratio.

### Correlations and agreements of QFR and UFR with FFR

Correlations and agreements among UFR, QFR, and FFR measurements were evaluated through correlation analysis and the Bland–Altman plots (Figure 2). The correlation of QFR and UFR with FFR was r = 0.68 (p < 0.001) and r = 0.79 (p < 0.001), respectively. The mean difference with FFR was -0.015 ± 0.08 (p = 0.006) for QFR and 0.001 ± 0.06 (p = 0.875) for UFR. These findings indicate significant correlations and agreements between both QFR and UFR with FFR. However, UFR demonstrated notably higher correlation with FFR compared to QFR (r = 0.79 versus 0.68, p = 0.011). Additionally, the Bland–Altman plots and ICC illustrated better agreement of FFR with UFR than with QFR (SD of the difference = 0.06 versus 0.08, p < 0.001; ICC = 0.79 [95% CI: 0.74-0.84] versus 0.67 [95% CI: 0.59-0.74], p = 0.007).

**Figure 2.**
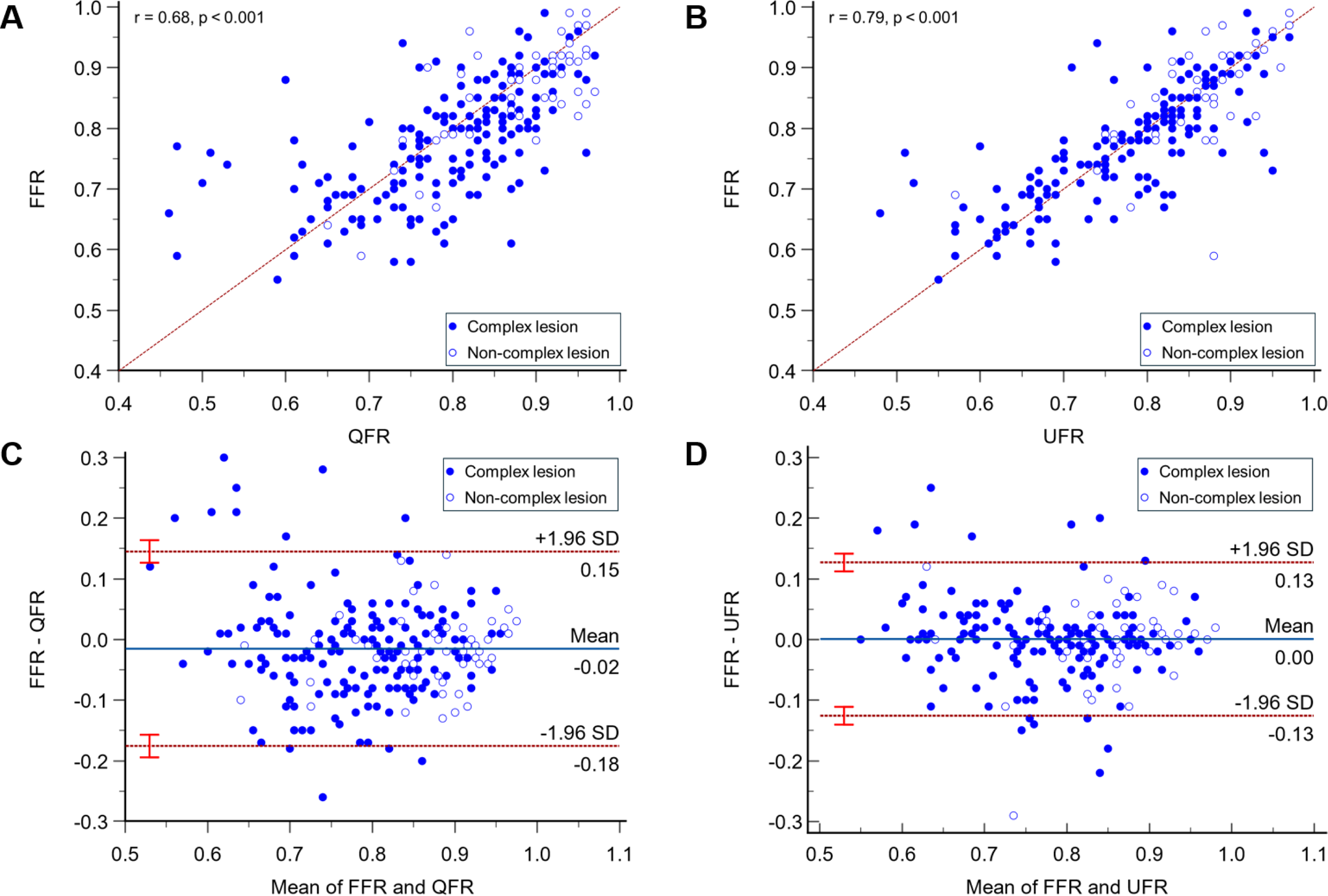
Correlation and agreement of QFR and UFR with FFR. **A** and **B**, Pearson correlations of QFR and UFR with FFR, respectively. **C** and **D**, Bland-Altman plots of QFR and UFR with FFR, respectively. QFR, quantitative flow reserve; UFR, ultrasonic flow ratio; FFR, fractional flow reserve.

### Diagnostic performances of QFR, UFR and IVUS-derived stenosis parameters

An arbitrary cut-off of 0.80 was used to define the physiological significance of stenosis, with values ≤0.80 for QFR, UFR and FFR indicating a physiologically significant lesion. The diagnostic performance of QFR and UFR in predicting FFR values is presented in Table 3. Discrepancies between FFR and QFR were found in 50 out of 220 vascular territories, while discrepancies between FFR and UFR were noted in 33 out of 220 territories. The diagnostic concordance between UFR and FFR (85.0% [95% CI: 79.6%-89.4%]) was significantly higher than that between QFR and FFR (77.3% [95% CI: 71.2%-82.6%], p=0.039). For QFR, the sensitivity and specificity in identifying patients with physiologically significant CAD were 68.1% and 87.5%, respectively. In comparison, UFR showed higher sensitivity and specificity at 80.2% and 90.4%, respectively. Additionally, in the ROC analysis (Figure 3A), UFR (AUC=0.91, [95% CI: 0.86–0.94]) outperformed QFR (AUC=0.86, [95% CI: 0.81–0.90]) in predicting the presence of lesions with FFR ≤0.80. The optimal cut-off values for IVUS-derived MLA and maximal plaque burden in predicting FFR ≤0.80 were determined to be 1.83 mm² and 77.3%, respectively.

**Figure 3.**
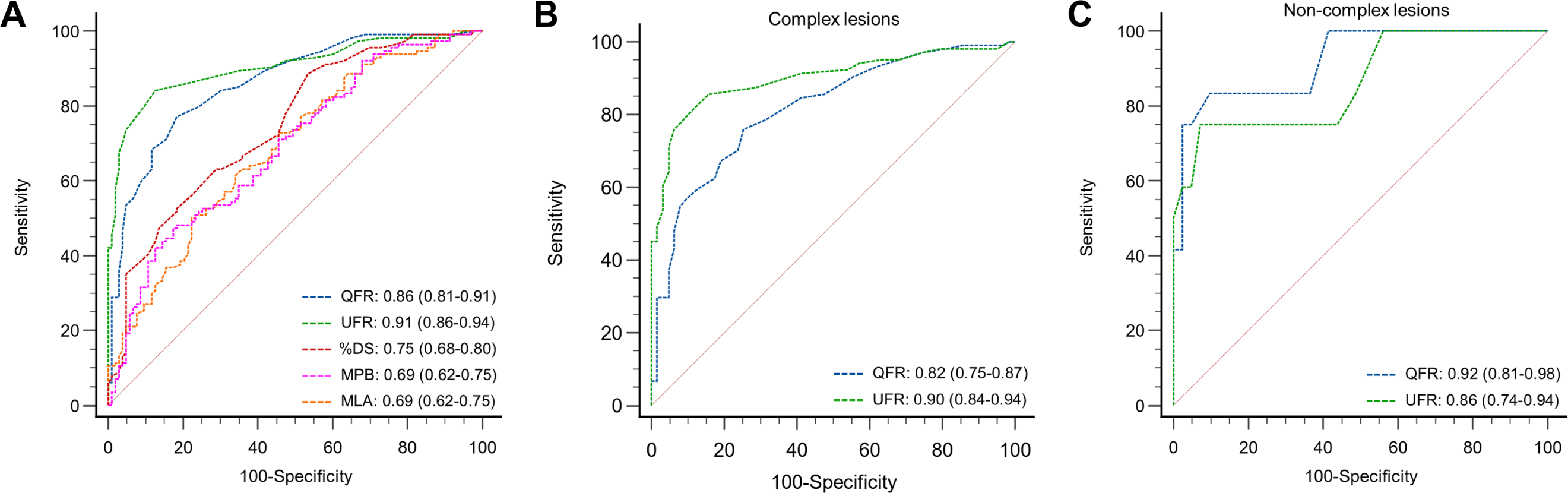
ROC curves for diagnosis of physiologically significant stenosis with FFR≤0.80. **A**, Comparison of diagnostic performance for QFR, UFR, IVUS-derived and MLA, and QCA-derived %DS. **B-C**, Comparison of diagnostic performance for QFR and UFR in complex and non-complex lesions. ROC, receiver operating characteristic; QFR, quantitative flow reserve; UFR, ultrasonic flow ratio; FFR, fractional flow reserve; IVUS, intravascular ultrasound; MLA, minimal lumen area; MPB, maximal plaque burden; QCA, quantitative coronary angiography; %DS, percent diameter stenosis.

**Table 3.** Diagnostic performances of UFR, QFR and IVUS and QCA-derived stenosis parameters.

|  | QFR<br>≤0.80 | UFR<br>≤0.80 | IVUS-derived MLA<br>≤1.83 | IVUS-derived MPB<br>>77.3% | QCA-derived %DS<br>>42.8% |
| --- | --- | --- | --- | --- | --- |
| Accuracy, % | 77.3 (71.2-82.6) | 85.0 (79.6-89.4) | 62.7 (56.0-69.1) | 63.6 (56.9-70.0) | 65.0 (58.3-71.3) |
| Sensitivity, % | 68.1 (58.8-76.4) | 80.2 (71.7-87.0) | 49.1 (39.7-58.6) | 47.4 (38.1-56.9) | 66.4 (57.0-74.9) |
| Specificity, % | 87.5 (79.6-93.2) | 90.4 (83.0-95.3) | 77.9 (68.7-85.4) | 81.7 (72.9-88.6) | 63.5 (53.4-72.7) |
| PPV, % | 85.9 (78.3-91.1) | 90.3 (83.7-94.4) | 71.3 (62.3-78.8) | 74.3 (64.9-81.9) | 67.0 (60.4-72.9) |
| NPV, % | 71.1(65.1-76.4) | 80.3 (73.8-85.6) | 57.9 (52.8-62.8) | 58.2 (53.4-62.9) | 62.9 (55.8-69.4) |
| +LR | 5.45 (3.23-9.20) | 8.34 (4.59-15.13) | 2.22 (1.48-3.33) | 2.60 (1.66-4.07) | 1.82 (1.37-2.42) |
| -LR | 0.37 (0.28-0.48) | 0.22 (0.15-0.32) | 0.65 (0.53-0.80) | 0.64 (0.53-0.78) | 0.53 (0.40-0.72) |
| AUC | 0.860 (0.807-0.903) | 0.906 (0.859-0.941) | 0.687 (0.620-0.748) | 0.686 (0.620-0.747) | 0.748 (0.685-0.804) |

QFR, quantitative flow reserve; UFR, ultrasonic flow ratio; IVUS, intravascular ultrasound; MLA, minimal lumen area; MPB, maximal plaque burden; QCA, quantitative coronary angiography; %DS, percent diameter stenosis; PPV, positive predictive value; NPV, negative predictive value; +LR, positive likelihood ratio; –LR, negative likelihood ratio; AUC, area under the curve.

### Diagnostic Performance of QFR and UFR in complex lesions

In this study, complex lesion subtypes included diffuse, bifurcation, and heavily calcified lesions. Lesions without these characteristics were classified as non-complex lesions (n=53, 24.1%). The diagnostic performance of QFR and UFR in the complex and non-complex lesions is summarized in Table 4. There was no significant difference in the diagnostic accuracy of UFR between complex and non-complex lesions (86.8% versus 84.4%, p=0.660). However, the presence of complex lesions significantly decreased the diagnostic accuracy of QFR (72.5% versus 92.5%, p=0.003). Additionally, in the ROC analysis (Figure 3B-C), the AUC for QFR was 0.82 (95% CI: 0.75–0.87) for complex lesions and 0.92 (95% CI: 0.81–0.98) for non-complex lesions. For UFR, the AUC was 0.90 (95% CI: 0.84–0.94) for complex lesions and 0.86 (95% CI: 0.74–0.94) for non-complex lesions. In complex lesions, the diagnostic concordance between UFR and FFR was significantly higher than that between QFR and FFR (86.8% versus 72.5%, p=0.001). Conversely, in non-complex lesions, the diagnostic concordance between UFR and FFR was numerically lower than that between QFR and FFR (84.4% versus 92.5%), though the difference was not statistically significant (p=0.194).

**Table 4.** Diagnostic performances of UFR and QFR in the complex and non-complex lesions.

|  | Complex lesions (n=167) |  | Non-complex lesions (n=53) |  |
| --- | --- | --- | --- | --- |
|  | QFR<br>≤0.80 | UFR<br>≤0.80 | QFR<br>≤0.80 | UFR<br>≤0.80 |
| Accuracy, % | 72.5 (65.0-79.1) | 84.4 (78.0-89.6) | 92.5 (81.8-97.9) | 86.8 (74.7-94.5) |
| Sensitivity, % | 67.3 (57.4-76.2) | 82.7 (74.0-89.4) | 75.0 (42.8-94.5) | 58.3 (27.7-84.8) |
| Specificity, % | 81.0 (69.1-89.8) | 87.3 (76.5-94.4) | 97.6 (87.1-99.9) | 95.1 (83.5-99.4) |
| PPV, % | 85.4 (77.5-90.8) | 91.5 (84.8-95.4) | 90.0 (55.8-98.5) | 77.8 (45.5-93.6) |
| NPV, % | 60.0(65.0-79.1) | 75.3 (66.5-82.5) | 93.0(83.3-97.3) | 88.6 (79.9-93.9) |
| +LR | 3.53 (2.09-5.98) | 6.51(3.39-12.52) | 30.75 (4.32-219.03) | 11.96 (2.85-50.16) |
| -LR | 0.40 (0.30-0.55) | 0.20 (0.13-0.31) | 0.26 (0.10-0.68) | 0.44 (0.22-0.86) |
| AUC | 0.817 (0.750-0.872) | 0.900 (0.844-0.941) | 0.921 (0.813-0.977) | 0.863 (0.740-0.942) |

QFR, quantitative flow reserve; UFR, ultrasonic flow ratio; PPV, positive predictive value; NPV, negative predictive value; +LR, positive likelihood ratio; –LR, negative likelihood ratio; AUC, area under the curve.

Further analysis was conducted to assess the impact of different complex lesion subtypes on the diagnostic performance of QFR and UFR (Figure 4). The diagnostic accuracy of QFR remained unaffected by the presence of diffuse lesions (AUC=0.86 vs. 0.84, p=0.718). However, UFR demonstrated a trend toward higher diagnostic performance in vessels with diffuse lesions compared to those without (AUC=0.92 vs. 0.84, p=0.126). UFR also showed similar diagnostic accuracy in vessels with and without bifurcation lesions (AUC=0.89 vs. 0.91, p=0.630). In contrast, the diagnostic performance of QFR was slightly reduced in vessels with bifurcation lesions compared to those without (AUC=0.78 vs. 0.89, p=0.059). Consistent with this, the presence of heavily calcified lesions significantly impaired the diagnostic performance of QFR (AUC=0.75 vs. 0.92, p=0.009), whereas it had no significant impact on UFR performance (AUC=0.88 vs. 0.91, p=0.543).

**Figure 4.**
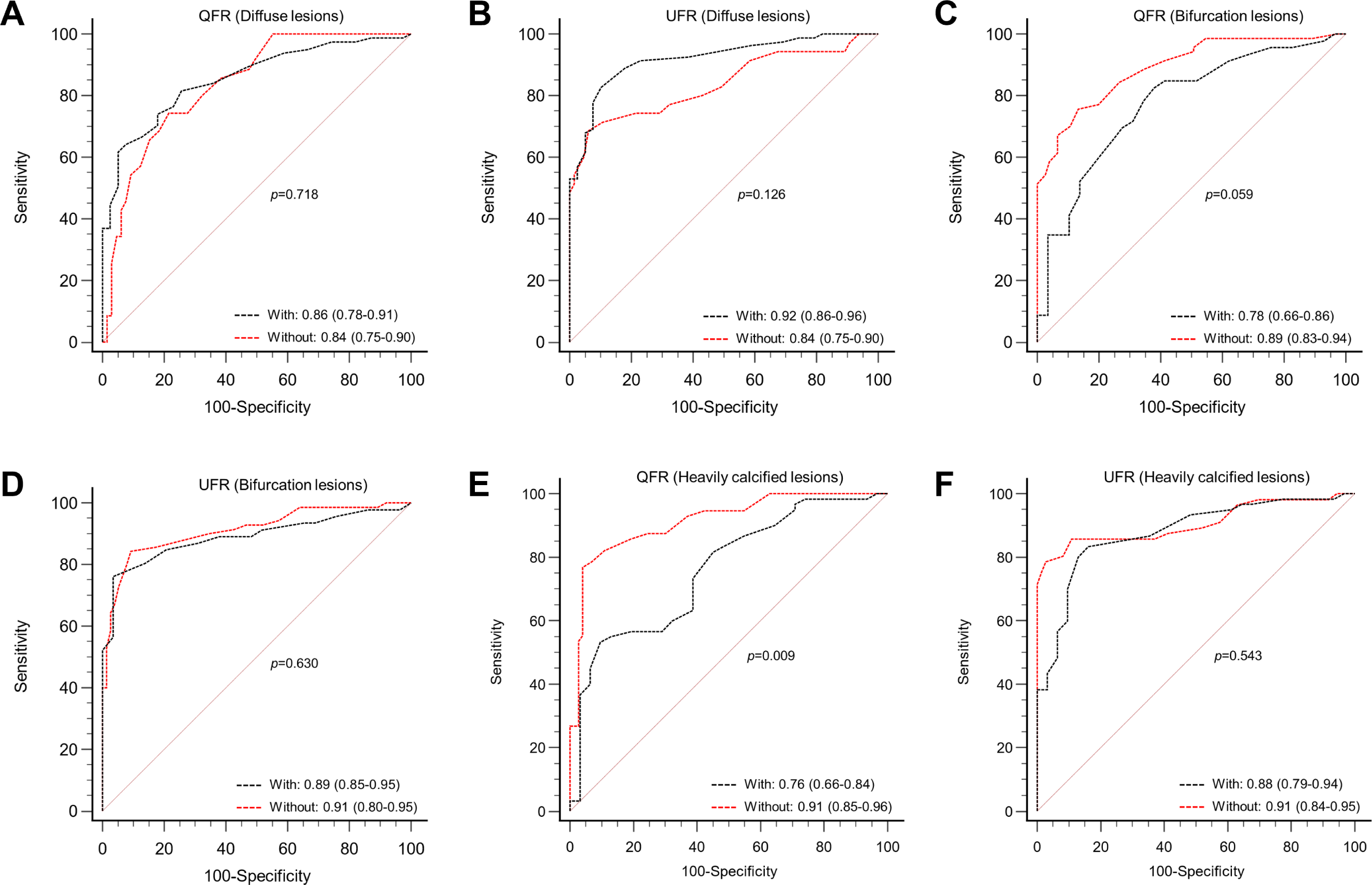
ROC curves analysis for the diagnostic values of QFR and UFR in different complex lesions. **A**, Diffuse lesions. **B**, Bifurcation lesions. **C**, Heavily calcified lesions. ROC, receiver operating characteristic; QFR, quantitative flow reserve; UFR, ultrasonic flow ratio.

### Diagnostic performances of QFR and UFR in the measurement grey zone

A total of 92 (41.8%) vessels had an FFR value falling within the measurement grey zone between 0.75 and 0.85. The diagnostic performance of UFR was better than that of QFR in the measurement grey zone, even though the difference was not statistically significant (AUC=0.77 versus 0.70, p=0.232). Accuracy, sensitivity, specificity, positive predictive value, negative predictive value, positive likelihood ratio and negative likelihood ratio for QFR were 63.0%, 45.8%, 81.8%, 73.3%, 58.1%, 2.5 and 0.7, and for UFR were 76.1%, 64.6%, 88.6%, 86.1%, 69.6%, 5.7 and 0.4, respectively.

## DISCUSSION

This study aimed to evaluate the diagnostic performance of UFR derived from intravascular IVUS and QFR derived from CAG in comparison with the gold standard FFR in patients with stable CAD. The main findings of our study are as follows: both UFR and QFR showed a significant correlation with FFR, highlighting their potential as reliable diagnostic tools for assessing hemodynamically significant coronary lesions . However, UFR showed superior diagnostic accuracy compared to QFR, particularly in complex coronary lesions, and its diagnostic superiority persisted even in the measurement grey zone (FFR 0.75–0.85). These findings support the utility of UFR as a reliable alternative for guiding clinical decision-making in complex coronary artery disease.

### Diagnostic performances of QFR and UFR

In our study, the diagnostic accuracy of QFR was notably lower than reported in previous literature. Prior studies suggested that QFR could achieve diagnostic accuracy rates exceeding 90% ^6,7^. However, in our findings, QFR showed a diagnostic concordance of 77.3% with FFR, with its accuracy further declining to 72.5% in complex lesions. This discrepancy can largely be attributed to the real-world nature of our patient cohort, which encompassed a high proportion of complex lesions (75.9%). The simultaneous use of FFR and IVUS generally indicates that the lesions were anatomically complex or ambiguous, where standard anatomical assessment alone would be insufficient to guide treatment decisions ^12,13^. Complex lesions can impair the quality of coronary angiography images, and in our study, only 37.3% of angiographic images met the criteria for optimum quality.

For example, calcified lesions often result in poor visualization of coronary artery edges, while bifurcation lesions may cause image overlap, affecting QFR calculation ^14^.

In contrast, UFR demonstrated consistently strong diagnostic performance across all lesion types, with an overall concordance of 85.0% and only a minimal drop in accuracy for complex lesions (86.8%). This consistency highlights UFR’s robustness in challenging anatomical contexts, which is a critical advantage in real-world clinical practice. Unlike QFR, UFR benefits from the detailed anatomical data provided by IVUS, allowing for more precise reconstruction of the vessel lumen and plaque burden, leading to more reliable estimation of hemodynamic significance ^8,15^.

### Comparison between QFR and UFR

Our results demonstrate that UFR offers superior diagnostic accuracy compared to QFR, especially in complex coronary lesions. While both methods rely on computational fluid dynamics to estimate the pressure gradient across a stenosis, UFR benefits from the three-dimensional imaging capability of IVUS, which allows for more accurate reconstruction of lesion geometry and better visualization of plaque morphology ^15,16^.

In complex scenarios like bifurcation and heavily calcified lesions, UFR maintained an accuracy of 86.8%, significantly higher than QFR’s 72.5%. QFR’s reliance on CAG makes it vulnerable to errors in scenarios with vessel overlap, foreshortening, and complex branching, which are common in bifurcations ^17,18^. Additionally, the presence of extensive calcification can impair the accuracy of QFR calculations ^18^, whereas UFR’s direct imaging approach is less susceptible to such issues. These findings suggest that UFR is a more robust option for functional assessment in patients with complex coronary anatomy.

### Clinical Implications

UFR’s integration with IVUS provides both anatomical and functional insights, which are particularly advantageous in managing complex coronary lesions ^12,19^. This dual-modality approach allows clinicians to make informed treatment decisions by accurately characterizing plaque and assessing vessel dimensions, thereby enhancing the precision of percutaneous coronary intervention (PCI) planning. For bifurcation and heavily calcified lesions, UFR provides critical evaluations that support effective stent placement and procedural planning, reducing the risk of suboptimal outcomes ^20^. Moreover, UFR avoids the need for pharmacological hyperemia required by FFR, making it a more patient-friendly option suitable for routine clinical use.

Given the high proportion of complex lesions in our study cohort, UFR’s ability to maintain high diagnostic accuracy underscores its value as a less invasive alternative to FFR. Its consistent diagnostic performance, even in the measurement grey zone, suggests that UFR could serve as an effective tool for guiding interventions, improving patient outcomes, and avoiding unnecessary procedures.

### Limitations

Despite the valuable insights provided by this study, several limitations should be acknowledged. First, the retrospective nature of the analysis may introduce selection bias, as patients included in the study were those who underwent both IVUS and FFR assessments. Additionally, the exclusion of certain patient groups, such as those with prior CABG, limits the generalizability of the findings to all complex CAD cases. Furthermore, the study relies on the accuracy of IVUS imaging for UFR calculation, and poor-quality IVUS images may lead to erroneous assessments. Finally, the study cohort was relatively small, and further prospective research with larger patient populations is needed to validate these findings and explore the full potential of UFR in different clinical settings.

## CONCLUSIONS

In summary, our study demonstrates that while both QFR and UFR are valuable tools for assessing coronary stenosis, UFR consistently outperforms QFR in terms of diagnostic accuracy, particularly in complex lesions such as diffuse, bifurcation, and heavily calcified lesions. By combining both anatomical and functional insights, UFR, when integrated with IVUS, provides a comprehensive and accurate assessment of coronary lesions, making it a powerful tool for guiding PCI. However, despite these promising results, further research with larger prospective cohorts is required to confirm these findings and refine the clinical applications of UFR.

## Data Availability

Data will be made available on request.

## ABBREVIATIONS

AUC: area under the curve
CABG: coronary artery bypass grafting
CAD: coronary artery disease
CAG: coronary angiography
FFR: fractional flow reserve
ICC: intraclass correlation coefficients
IVUS: intravascular ultrasound
MLA: minimum lumen area
PCI: percutaneous coronary intervention
QCA: quantitative coronary angiography
QFR: quantitative flow ratio
ROC: receiver operating characteristic
UFR: ultrasonic flow ratio.

